# Anatomy-guided, modality-agnostic segmentation of neuroimaging abnormalities

**DOI:** 10.1101/2025.04.29.25326682

**Authors:** Diala Lteif, Divya Appapogu, Sarah A. Bargal, Bryan A. Plummer, Vijaya B. Kolachalama

## Abstract

Magnetic resonance imaging (MRI) offers multiple sequences that provide complementary views of brain anatomy and pathology. However, real-world datasets often exhibit variability in sequence availability due to clinical and logistical constraints. This variability complicates radiological interpretation and limits the generalizability of machine learning models that depend on consistent multimodal input. In this work, we propose an anatomy-guided and modality-agnostic framework for assessing disease-related abnormalities in brain MRI, leveraging structural context to enhance robustness across diverse input configurations. We introduce a novel augmentation strategy, Region ModalMix, which integrates anatomical priors during training to improve model performance when some modalities are absent or variable. We conducted extensive experiments on brain tumor segmentation using the Multimodal Brain Tumor Segmentation Challenge (BraTS) 2020 dataset (n=369). The results demonstrate that our proposed framework outperforms state-of-the-art methods on various missing modality conditions, especially by an average 9.68 mm reduction in 95^th^ percentile Hausdorff Distance and a 1.36% improvement in Dice Similarity Coefficient over baseline models with only one available modailty. Our method is model-agnostic, training-compatible, and broadly applicable to multi-modal neuroimaging pipelines, enabling more reliable abnormality detection in settings with heterogeneous data availability.

## 1 Introduction

Magnetic resonance imaging (MRI) is a cornerstone of neuroradiology, offering multiple sequences such as T1-weighted and fluid-attenuated inversion recovery (FLAIR) that provide complementary information for assessing brain structure and pathology. These multimodal sequences are critical for identifying and differentiating conditions such as ischemic strokes, tumors, and neurodegenerative disorders. However, in both clinical and research settings, acquiring complete multimodal MRI data is often constrained by time, cost, computational demands, and patient tolerance. As a result, many datasets lack one or more sequences, limiting their diagnostic value and complicating the development of robust machine learning (ML) models [31, 15]. In clinical environments, imaging protocols may be abbreviated to prioritize urgent care, while in research, cohort studies frequently include only a subset of modalities due to logistical or financial limitations. These gaps reduce the utility of otherwise valuable datasets and pose a challenge for multimodal model training and deployment. Nonetheless, developing methods that can leverage incomplete imaging data could unlock the full potential of these datasets, advancing clinical applications and generalizable ML research in neuroimaging.

Traditional approaches to handling incomplete multi-modal data typically train ML models on fully observed datasets and simulate missingness only at evaluation time [5, 28, 24, 27]. An alternative line of work aims to synthesize missing sequences [7, 11, 25, 4], but such generative models often require large, fully-sampled datasets and may not capture the full spectrum of clinical variability. Moreover, many segmentation frameworks trained on modality-complete inputs are brittle to missing sequences, resulting in performance degradation when applied to modality-incomplete data.

To address these limitations, we propose *Region ModalMix* (RMM), a data augmentation strategy designed to improve the robustness of state-of-the-art brain tumor segmentation models even under conditions of modality incompleteness. Our method leverages anatomical priors in the form of parcellation maps to guide the mixing of available modalities within predefined brain regions. By augmenting the training process with anatomically coherent, cross-modal representations derived from the same subject, RMM promotes resilience to missing modalities without requiring explicit imputation or synthesis. This framework enhances model generalizability, improves segmentation accuracy in incomplete data scenarios, and expands the applicability of high-performing ML models to real-world neuroimaging datasets.

## 2 Related work

Several studies have investigated unified models for tasks such as brain tumor segmentation from incomplete MRI modalities [28, 5, 2, 23, 19, 8, 14, 6, 26, 9]. One line of research focuses on multimodal fusion using various attention mechanisms [5, 30, 12]. For instance, the Region-aware Fusion Network (RFNet [5]) employed a 3D U-Net [3] architecture with separate modality-specific convolutional encoders, a shared-weight decoder used for regularization, and a late-stage fusion decoder utilizing their proposed Region-aware Fusion Module (RFM). RFM adaptively fuses available modal features based on tumor regions (i.e., whole tumor (WT), tumor core (TC), and enhancing tumor (ET)), thereby augmenting segmentation performance across various modality combinations.

Some approaches have adapted transformers for incomplete multimodal learning [28, 24, 27]. The multimodal medical transformer (mmFormer [28]) was the first of those approaches to incorporate hybrid modality-specific transformer-based encoders alongside a shared modality-correlated transformer-based encoder, facilitating the learning of modality-specific and cross-modal features. However, maintaining separate encoders for each modality can incur computational overhead [27]. To address this, IMS^2^Trans [27] introduced a lightweight architecture with a single encoder for all modalities, reducing redundancy while effectively handling missing modalities. Similarly, TMFormer [29] merged available modalities into compact tokens, streamlining the fusion process. While these proposed architectures adopted vanilla pairwise self-attention, others (e.g., M^2^FTrans [24]) introduced alternative solutions using modality-masked fusion strategies with learnable fusion tokens and spatial weight attention, to adaptively re-weight modalities for better fusion. Despite their efficiency, these methods do not take advantage of the prior knowledge available for brain MRI datasets, which could provide a wealth of insight into the training process, yielding clinically relevant models. In our work, we used the anatomical context of the human brain to learn robust representations of incomplete multimodal data, even when training data was incomplete. Our framework was designed to be model-agnostic and offers flexibility across various architectures.

## 3. Methods

### 3.1 Study population and data processing

We conducted experiments on the task of modality-incomplete brain tumor segmentation using the BraTS 2020 dataset. The cohort was compiled from 19 institutions as part of the Multimodal Brain Tumor Segmentation Challenge [17] and includes 369 subjects, each with four MRI sequences: T1-weighted, T2-weighted, T1 contrast-enhanced (T1ce), and FLAIR. Voxel-wise annotations are provided for three tumor subregions: nectoric and non-enhancing tumor (NCR/NET), edema (ED), and enhancing tumor (ET), enabling fine-grained segmentation of glioblastoma (HGG, *n* = 293) and lower-grade glioma (LGG, n=76) cases. Demographic details of the cohort are summarized in Table 1.

**Table 1:** Summary of the BraTS 2020 brain tumor segmentation dataset. This table summarizes participant demographics and MRI acquisition details for the dataset used in this study. The dataset is divided into two tumor grade cohorts: high-grade gliomas (HGG) and low-grade gliomas (LGG). The number of participants, total MRIs across four modalities (T2, T1ce, T1, FLAIR), and mean age with standard deviation are reported. Age data for LGG cases was not available (N.A.).

| Cohort<br>(group) | Participants<br>n (%) | MRIs<br>n (T2,T1ce,T1,F) | Age (y)<br>mean±s.d. |
| --- | --- | --- | --- |
| HGG | 293 | 1,172 | 61.22±11.87 |
| LGG | 76 | 304 | N.A. |

The BraTS dataset was provided in NIFTI format following a series of preprocessing steps, including skull stripping, co-registration to the SRI24 multi-channel anatomical template [21], and isotropic resampling to a resolution of 1 × 1 × 1 mm^3^. We converted the data into `numpy` format, removed non-brain regions, and normalized each modality to zero mean and unit variance, following protocols established by prior work [5]. Each scan was parcellated using SynthSeg+ [1], yielding volumetric labels for 99 distinct brain regions, along with cerebrospinal fluid and background, resulting in a total of 101 parcellated regions. These anatomical parcellations served as a foundation for our data augmentation strategy (Supplementary Table A1). Each multimodal sample was represented as a 4D tensor of shape *M* × *D* × *H* × *W*, where *M* is the number of modalities, and *D, H, W* represent the scan dimensions. The corresponding segmentation maps *Seg*^*m*^ of size *D* × *H* × *W* were used to guide the training of our models.

### 3.2 Modeling framework

We propose *Region ModalMix* (RMM), a novel data augmentation strategy desgined to enhance brain tumor segmentation performance in the presence of modality-incomplete MRI. As illustrated in Figure 1, our end-to-end framework integrates RMM into a multimodal segmentation pipeline to generate anatomically coherent and modality-diverse training samples. RMM leverages modality-specific brain parcellation maps to guide augmentation at the region level. The implementation details of RMM are outlined in Algorithm 1, where *B, M, N*, and *R* denote the batch size, the number of MRI modalities, the size of the training dataset, and the total number of brain regions (*R* = 99), respectively. The core process of RMM, described in lines 3 to 11 of Algorithm 1, operates as follows: at each training iteration, a batch of data (*X, Y, Seg*) is sampled from the training set, and a series of spatial and structural transformations is applied. To maintain uniformity of transformations across modalities and volumetric segmentations for the same participant, we employed a seeded transformation function. This function ensures consistent random transformations by fixing the random generator seed in `torch`, `monai`, and other relevant libraries. After applying the transformations, RMM generates new data 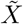 by randomly mixing brain regions along the modality dimension, i.e., the second tensor dimension. Using the `Seg` tensors, for a given modality *m*, pixels corresponding to an anatomical region *r* are replaced with those from the same region *r* in a randomly sampled modality *rm*. An example augmentation can be viewed in Figure 2, where mixed regions in the sample augmented by our method (b) are color-coded by their modality of origin. Notably, the target segmentation labels *Y* are not altered, as the mixing is performed across modalities of the same subject, and there is only one reference segmentation label corresponding to each of the samples, regardless of their multimodal nature.

**Figure 1:**
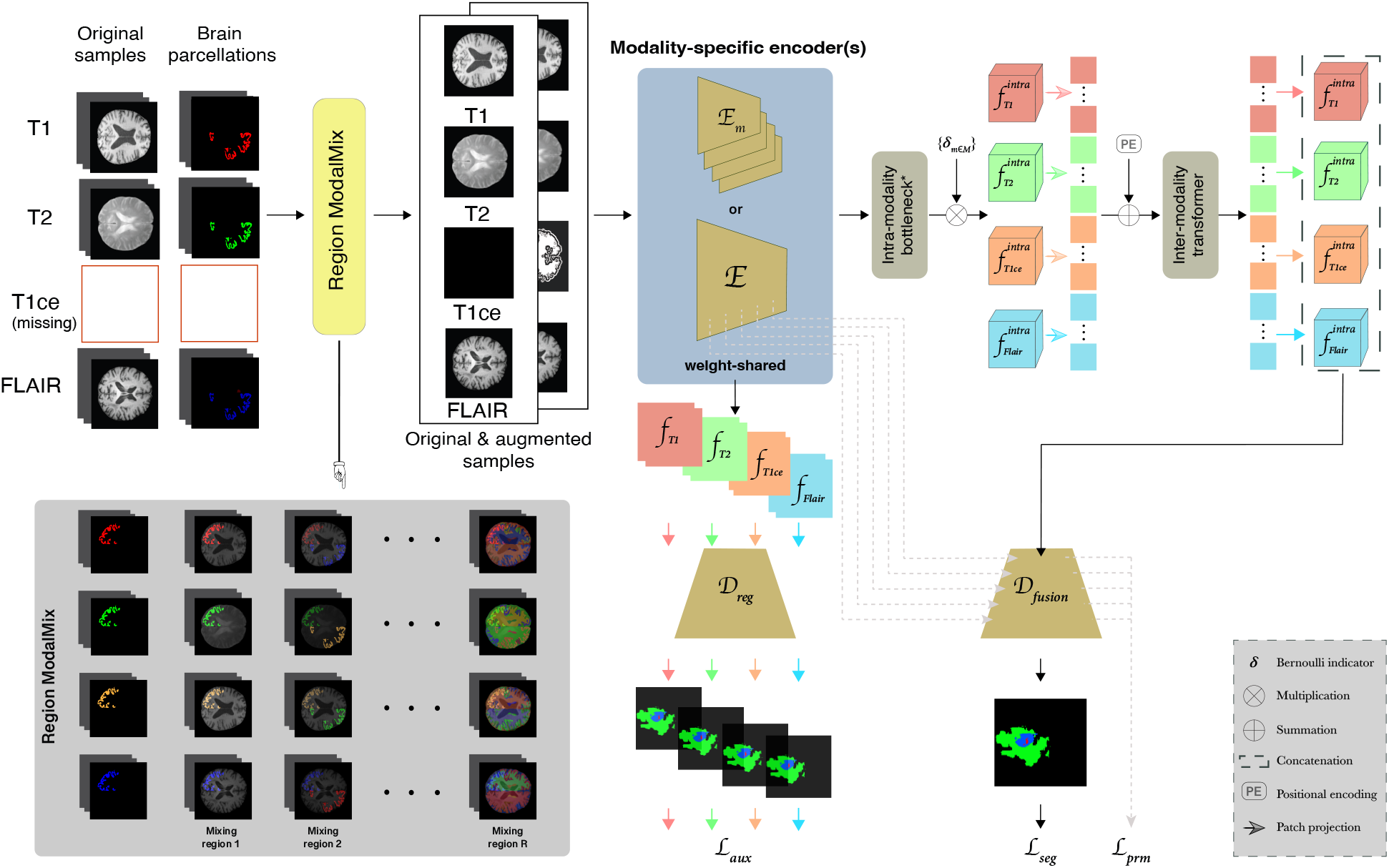
Overview of our framework. We introduce *Region ModalMix*, a data augmentation strategy that leverages approximately 100 brain parcellation masks to mix information across available MRI modalities (e.g., T1, T2, FLAIR) within predefined anatomical regions. During training, we randomly sample regions and mix them across modalities for the same subject, generating augmented inputs that are concatenated with the original data. These samples are processed by either a set of modality-specific encoder {*E*_*m*∈*M*_}, where *M* denotes the set of modalities, or a weight-shared encoder. The extracted features are passed through an intra-modality bottleneck and intermodality transformer backbone, followed by a decoder module *D*_*fusion*_. To enhance representation learning under missing modalities, we employ auxiliary regularization based on modality-specific predictions from *D*_*reg*_. Note that the intra-modality bottleneck may be either convolutional or transformer-based, depending on the architecture used. The procedure is outlined in Algorithm 1.

**Figure 2:**
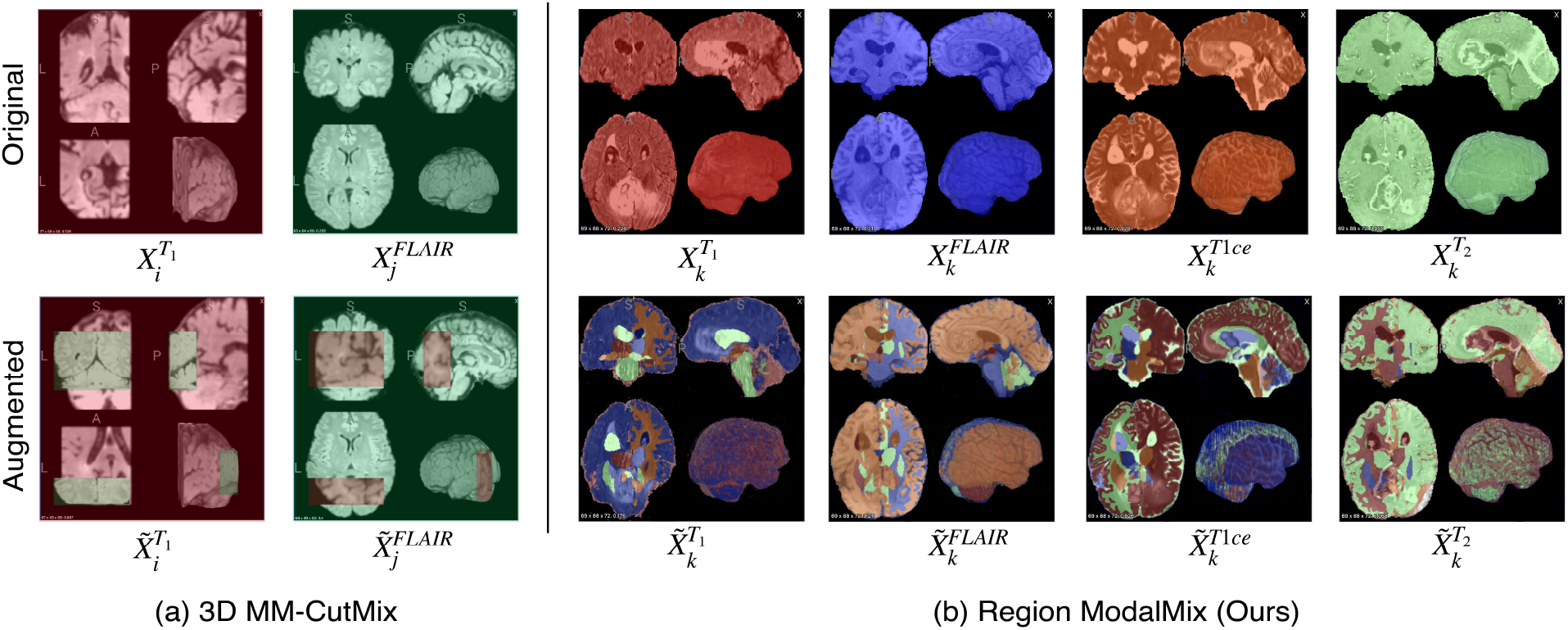
Comparison of 3D MM-CutMix and Region ModalMix (RMM). Examples of augmented samples generated by (a) 3D MM-CutMix and (b) our proposed RMM. 3D MM-CutMix combines cuboid patches from MRI scans of different participants and modalities. In contrast, RMM operates on scans from a single participant across multiple modalities, mixing anatomically defined regions to generate fine-grained, modality-aware augmentations.

#### Algorithm 1

Pseudocode for Region ModalMix

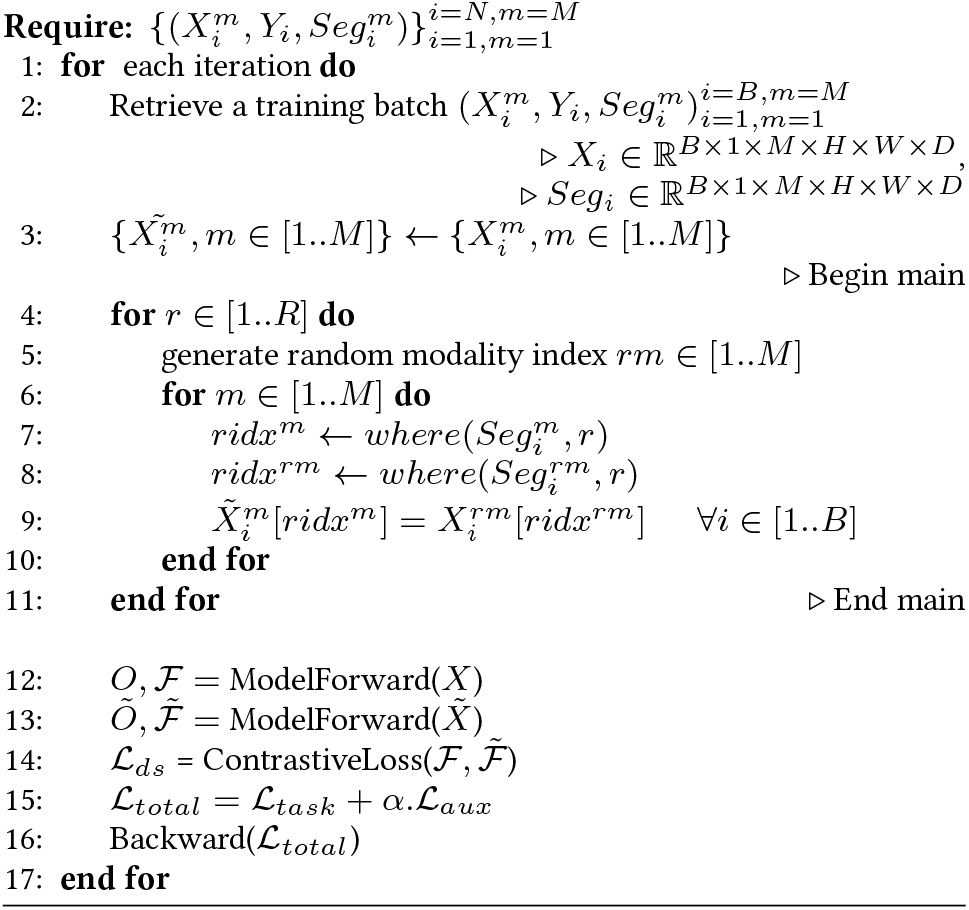

Unlike 3D MM-CutMix [27], which mixes cuboid patches across participants and modalities, RMM preserves anatomical plausibility by mixing only within the spatial context of a single subject. As illustrated in Figure 2, 3D MM-CutMix (a) introduces anatomically incoherent boundaries by stitching together unrelated volumes, whereas RMM (b) maintains structural continuity across regions. Once augmented, the mixed input is concatenated with the original data and passed through either a set of modality-specific encoders {*E*_*m*∈*M*_} or a shared encoder, depending on the model architecture. The encoded features are processed through an intra-modality bottleneck (either convolutional or transformer-based), followed by an inter-modality transformer module. A decoder head *D*_*fusion*_ then produces the final segmentation output. To enhance model performance under missing modality conditions, we incorporated an auxiliary regularization module with a corresponding loss term, ℒ_*aux*_, defined differently based on the underlying architecture. In models with modality-specific encoders and a shared-weight decoder [5, 28, 24], ℒ_*aux*_ was used to ensure that each modality-specific encoder learned discriminative features, even in the absence of certain modalities. Specifically, it was computed as:

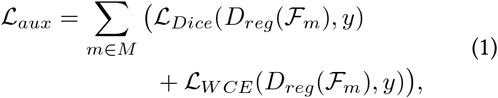

where *D*_*reg*_ denotes the shared-weight decoder used for regularization, ℱ_*m*_ is the feature representation extracted from the *m*-th modality-specific encoder, and *y* is the segmentation label. In IMS^2^Trans [27], ℒ _*aux*_ was implemented as a feature distillation loss via contrastive learning to align individual modality features with their mean feature representation. The overall training objective combined the primary task loss with the auxiliary loss:

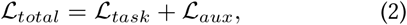

where ℒ_*task*_ was tailored to the segmentation task, particularly defined as:

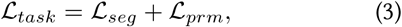

where ℒ_*seg*_ is the final segmentation loss and ℒ_*prm*_ is a deep supervision loss applied to intermediate outputs of the fusion decoder, encouraging consistent predictions across network depths. ℒ_*seg*_ was computed as a weighted combination of the Dice Similarity Coefficient (DSC) [18] and the Weighted Cross-Entropy (WCE) [22] loss functions. Additional details regarding the implementation can be found in the literature [5, 28, 24, 27].

**Figure 3:**
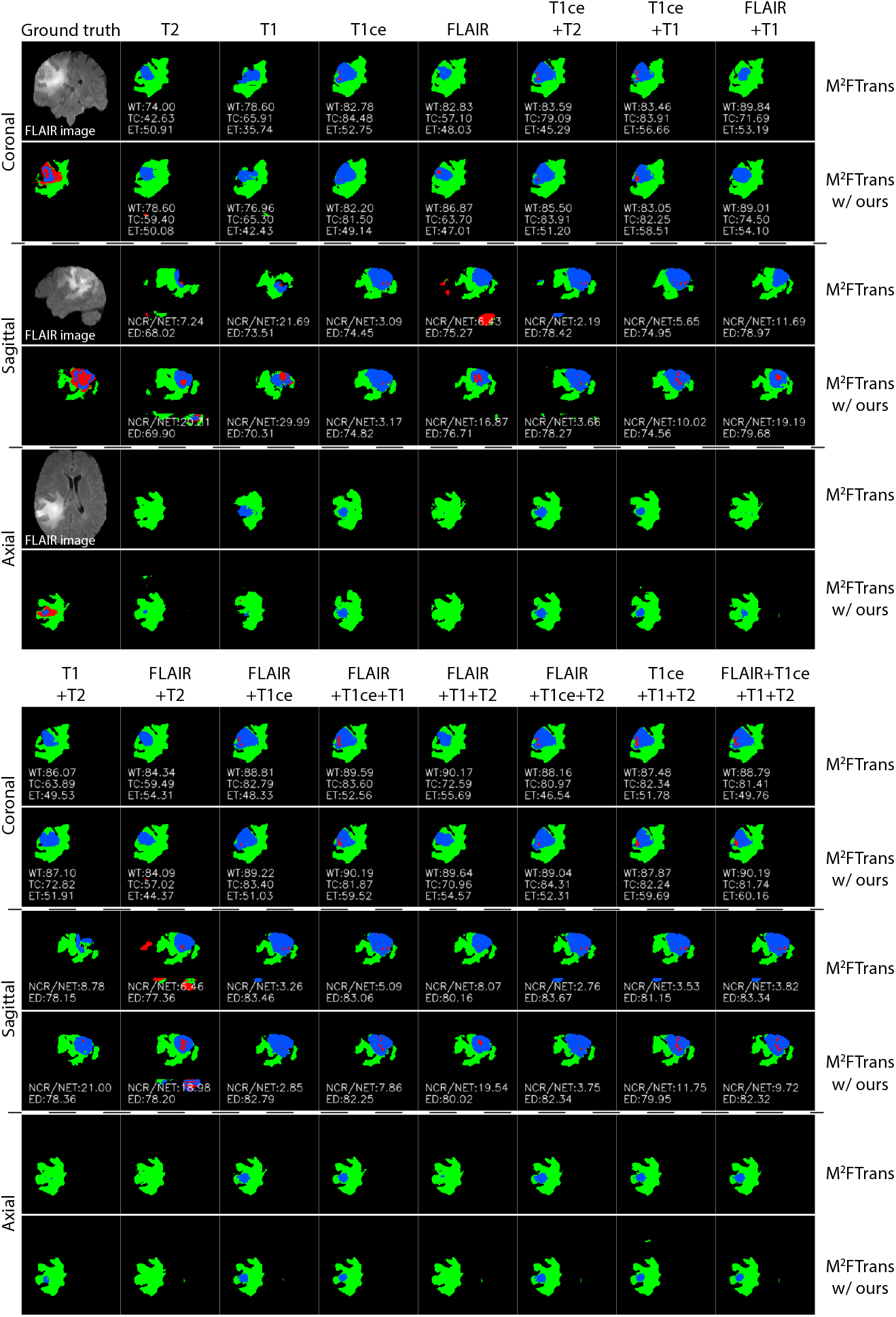
Qualitative segmentation comparison across modality combinations on BraTS 2020. Visualization of tumor segmentation results using M^2^FTrans [24] with and without our proposed augmentation strategy under 15 different MRI modality combinations. Each column represents a unique subset of available modalities. For each setting, we display the model’s predictions alongside ground truth segmentations, overlaid on FLAIR slices in axial, coronal, and sagittal views. Tumor subregions are color-coded: necrotic/non-enhancing tumor (NCR/NET) in red, edema (ED) in green, and enhancing tumor (ET) in blue. DSC scores (%) for each tumor region are shown within the corresponding coronal slice. Sagittal views also include region-specific DSC scores for finer-grained evaluation.

## 4 Experimental setup

### 4.1 Training and model settings

We evaluated our model on the task of brain tumor segmentation, comparing it against benchmark methods designed to handle modality-incomplete MRI data: RFNet [5], mmFormer [28], M^2^FTrans [24], and IMS^2^Trans [27]. Experiments were conducted on the BraTS 2020 dataset, adopted from the Brain Tumor Segmentation (BraTS) Challenge [17], which includes four MRI modalities: FLAIR, T1 contrast-enhanced (T1ce), T1-weighted, and T2-weighted, available for every subject. Following [5], we removed non-brain background regions and normalized each modality to have zero mean and unit variance. To ensure a fair comparison with state-of-the-art models, we strictly followed the data split protocol from [5, 28, 24, 27], dividing the 369 subjects into 219 for training, 50 for validation, and 100 for testing. Although BraTS 2020 is modality-complete, we simulated missing modality scenarios during training by randomly masking MRI sequences. Input volumes were randomly cropped to 64 × 64 × 64 voxels and augmented with random rotations, intensity shifts, and mirror flips.

All models were trained using a batch size of 2, an initial learning rate of 2e − 4, and a weight decay of 1e − 4 for 1000 epochs with 200 iterations per epoch. Training was conducted on a single NVIDIA GPU with 48GB memory. We used the AdamW [16] optimizer under identical experimental conditions, with preserved latent dimensionality and parameter count across all models. Refer to Supplementary Table A2 for details about model settings.

### 4.2 Performance metrics

Evaluation metrics for segmentation included the Dice Similarity Coefficient (DSC) [18], also known as the F1-score or Sørensen-Dice index, and the 95^th^ percentile Hausdorff Distance (HD95) [10]. DSC measures the global overlap between predicted and ground truth segmentations, but is less sensitive to local errors. HD95 measures the 95^th^ percentile of all the distances from a point of one boundary to the closest point on the other boundary. It complements DSC by capturing boundary-based deviations while being robust to outlier voxels, making it well-suited for medical image segmentation.

Following [5, 28, 24, 27], the tumor regions (i.e., NCR/NET, ED, and ET), were aggregated into three tumor classes for evaluation: whole tumor (WT), consisting of all three regions, tumor core (TC), comprising the NCR/NET and ET regions, and finally the ET region as a separate class. Performance was evaluated on the three tumor classes (i.e., WT, TC, and ET) as well as the NCR/NET and ED regions. The models were tested on 15 different modality combinations and comparisons were stratified by the number of missing modalities. For a fair comparison, all models were evaluated at the 1000^th^ epoch.

## 5. Results

Quantitative results on the BraTS 2020 dataset are summarized in Tables 2 and 3 showing average performance improvements over state-of-the-art methods, measured in HD95 (mm) and DSC (%) scores, respectively, under varying degrees of modality incompleteness. Across both HD95 (Table 2) and DSC (Table 3), RMM yielded consistent performance improvements, particularly when integrated with the M^2^FTrans back-bone [24]. For instance, M^2^FTrans [24] trained with our method showed the largest average HD95 reduction in multiple subregions—including WT (− 10.24 mm), TC (− 20.79 mm), and ET (− 20.23 mm) with no missing modalities—and consistently strong improvements under partial modality settings. These boundary-level improvements were mirrored in DSC gains, where the same model achieved the highest improvements in ET (+8.88%) and TC (+4.58%) when all modalities were available. These findings suggest that RMM not only enhances robustness in modality-incomplete scenarios but also boosts model performance when full information is available, likely due to its ability to promote better generalization via anatomically grounded augmentation.

**Table 2:**
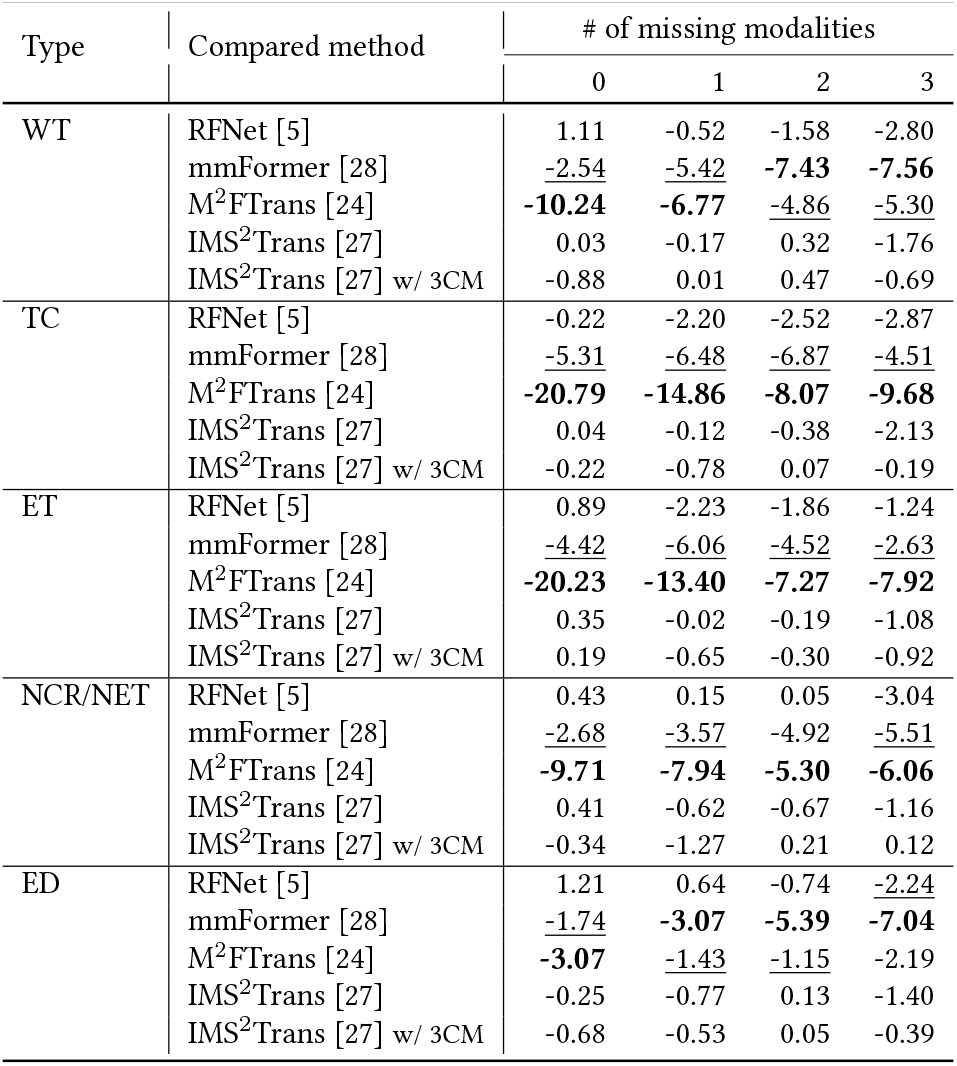
Performance gains (HD95, mm) of our framework over state-of-the-art methods under varying numbers of missing MRI modalities. This table reports the average improvements in segmentation accuracy, measured by the 95th percentile Hausdorff Distance (HD95), across different tissue types as the number of missing modalities increases. Negative values indicate improved performance (i.e., lower HD95) relative to the original method. For each setting, the best-performing improvement is shown in **bold**, and the second-best is <u>underlined</u>, where applicable.

| Type | Compared method | # of missing modalities |  |  |  |
| --- | --- | --- | --- | --- | --- |
|  |  | 0 | 1 | 2 | 3 |
| WT | RFNet [5] | 1.11 | -0.52 | -1.58 | -2.80 |
|  | mmFormer [28] | <u>-2.54</u> | <u>-5.42</u> | **-7.43** | **-7.56** |
|  | M<sup>2</sup>FTrans [24] | **-10.24** | **-6.77** | <u>-4.86</u> | -5.30 |
|  | IMS<sup>2</sup>Trans [27] | 0.03 | -0.17 | 0.32 | -1.76 |
|  | IMS<sup>2</sup>Trans [27] w/ 3CM | -0.88 | 0.01 | 0.47 | -0.69 |
| TC | RFNet [5] | -0.22 | -2.20 | -2.52 | -2.87 |
|  | mmFormer [28] | <u>-5.31</u> | <u>-6.48</u> | <u>-6.87</u> | <u>-4.51</u> |
|  | M<sup>2</sup>FTrans [24] | **-20.79** | **-14.86** | **-8.07** | **-9.68** |
|  | IMS<sup>2</sup>Trans [27] | 0.04 | -0.12 | -0.38 | -2.13 |
|  | IMS<sup>2</sup>Trans [27] w/ 3CM | -0.22 | -0.78 | 0.07 | -0.19 |
| ET | RFNet [5] | 0.89 | -2.23 | -1.86 | -1.24 |
|  | mmFormer [28] | <u>-4.42</u> | <u>-6.06</u> | <u>-4.52</u> | <u>-2.63</u> |
|  | M<sup>2</sup>FTrans [24] | **-20.23** | **-13.40** | **-7.27** | **-7.92** |
|  | IMS<sup>2</sup>Trans [27] | 0.35 | -0.02 | -0.19 | -1.08 |
|  | IMS<sup>2</sup>Trans [27] w/ 3CM | 0.19 | -0.65 | -0.30 | -0.92 |
| NCR/NET | RFNet [5] | 0.43 | 0.15 | 0.05 | -3.04 |
|  | mmFormer [28] | -2.68 | -3.57 | -4.92 | -5.51 |
|  | M<sup>2</sup>FTrans [24] | **-9.71** | **-7.94** | **-5.30** | **-6.06** |
|  | IMS<sup>2</sup>Trans [27] | 0.41 | -0.62 | -0.67 | -1.16 |
|  | IMS<sup>2</sup>Trans [27] w/ 3CM | -0.34 | -1.27 | 0.21 | 0.12 |
| ED | RFNet [5] | 1.21 | 0.64 | -0.74 | -2.24 |
|  | mmFormer [28] | <u>-1.74</u> | **-3.07** | **-5.39** | **-7.04** |
|  | M<sup>2</sup>FTrans [24] | **-3.07** | <u>-1.43</u> | <u>-1.15</u> | -2.19 |
|  | IMS<sup>2</sup>Trans [27] | -0.25 | -0.77 | 0.13 | -1.40 |
|  | IMS<sup>2</sup>Trans [27] w/ 3CM | -0.68 | -0.53 | 0.05 | -0.39 |

**Table 3:** Improvement in segmentation accuracy (DSC, %) across varying levels of modality availability. This table summarizes the average gain in Dice Similarity Coefficient (DSC) achieved by our framework compared to several state-of-the-art models under different numbers of missing MRI modalities. Positive values indicate performance gains, measured in percentage points. For each scenario, the highest improvement is highlighted in **bold** and the second highest is <u>underlined</u>, where applicable. Results are shown separately for different tissue types.

| Type | Compared method | # of missing modalities |  |  |  |
| --- | --- | --- | --- | --- | --- |
|  |  | 0 | 1 | 2 | 3 |
| WT | RFNet [5] | -0.93 | -0.71 | -0.88 | -1.18 |
|  | mmFormer [28] | 0.33 | <u>0.78</u> | **1.09** | **1.36** |
|  | M<sup>2</sup>FTrans [24] | **1.74** | **1.31** | <u>1.02</u> | <u>0.44</u> |
|  | IMS<sup>2</sup>Trans [27] | -0.02 | 0.11 | -0.04 | -0.51 |
|  | IMS<sup>2</sup>Trans [27] w/ 3CM | <u>0.82</u> | 0.64 | 0.23 | -0.67 |
| TC | RFNet [5] | 0.06 | -0.32 | -1.12 | -3.13 |
|  | mmFormer [28] | <u>2.16</u> | <u>1.73</u> | <u>0.55</u> | -2.30 |
|  | M<sup>2</sup>FTrans [24] | **4.58** | **3.00** | **1.36** | -0.31 |
|  | IMS<sup>2</sup>Trans [27] | -0.67 | 0.06 | -0.75 | -4.28 |
|  | IMS<sup>2</sup>Trans [27] w/ 3CM | 0.19 | 0.38 | -0.84 | -4.37 |
| ET | RFNet [5] | 1.00 | 0.13 | 1.16 | **1.15** |
|  | mmFormer [28] | <u>6.24</u> | 2.42 | 0.75 | -2.04 |
|  | M<sup>2</sup>FTrans [24] | **8.88** | **4.53** | 1.29 | -0.88 |
|  | IMS<sup>2</sup>Trans [27] | 2.58 | 2.34 | <u>1.51</u> | -1.03 |
|  | IMS<sup>2</sup>Trans [27] w/ 3CM | 2.90 | <u>2.73</u> | **2.79** | -1.02 |
| NCR/NET | RFNet [5] | -1.08 | -1.70 | -1.95 | -2.51 |
|  | mmFormer [28] | <u>1.87</u> | **1.57** | <u>0.44</u> | -0.90 |
|  | M<sup>2</sup>FTrans [24] | **1.91** | 1.52 | **0.98** | **0.20** |
|  | IMS<sup>2</sup>Trans [27] | 0.06 | 0.31 | -0.72 | -2.75 |
|  | IMS<sup>2</sup>Trans [27] w/ 3CM | 0.56 | 0.28 | -1.07 | -3.11 |
| ED | RFNet [5] | -0.72 | -0.89 | -1.08 | -1.11 |
|  | mmFormer [28] | 0.83 | <u>0.72</u> | **0.82** | **1.26** |
|  | M<sup>2</sup>FTrans [24] | <u>0.99</u> | **0.77** | <u>0.49</u> | -0.10 |
|  | IMS<sup>2</sup>Trans [27] | 0.43 | 0.38 | 0.33 | <u>0.06</u> |
|  | IMS<sup>2</sup>Trans [27] w/ 3CM | **1.05** | 0.66 | 0.35 | -0.18 |

Importantly, ET and NCR/NET are among the smallest and most spatially complex tumor subregions, posing significant challenges for accurate segmentation. The results in Tables 2 and 3 demonstrate that our method substantially improves HD95 and DSC in these regions across models. For example, in ET, RMM integrated with the M^2^FTrans [24] backbone achieved a − 20.23 mm HD95 improvement and a +8.88% DSC gain, while in NCR/NET, we see consistent HD95 reductions and DSC gains across all levels of missingness. This indicates that the anatomical focus of RMM helps improve spatial delineation and volume estimation in fine-grained structures, which are often underrepresented in standard training settings.

To our knowledge, we are the first to provide detailed performance evaluations for Edema (ED) and NCR/NET in a modality-incomplete setup—regions typically omitted from prior multimodal segmentation benchmarks. Despite this, RMM achieves robust improvements in both regions—e.g., mmFormer trained with RMM yielded a *−* 7.04 mm HD95 reduction and a +1.26% DSC gain in ED under the most severe missingness condition. Detailed results in Supplementary Tables A3 and A4 further corroborated these findings. Although the DSC metric is known to be sensitive to structure size, topology, and disconnectedness [20], especially for regions like NCR/NET, RMM improved performance across multiple models (e.g., mmFormer [28] and M^2^FTrans [24]) on both NCR/NET and ED. As for HD95, a boundary-aware metric that is more suitable for finer-grained structures, our method showed substantial reductions in both subregions across all backbones. This emphasizes our method’s ability to generalize beyond the conventional WT/TC/ET segmentation targets and extend to anatomically diverse and clinically relevant subregions.

RMM also outperforms 3D MM-CutMix, which served as the most relevant prior data augmentation method proposed for brain tumor segmentation. Although 3CM provides modest benefits as evident in Supplementary Tables A3 and A4, RMM consistently achieved higher improvements across both metrics (e.g., a 2.79% DSC gain in ET with two missing modalities). Even models with strong baseline performance, such as RFNet [5] (see Supplementary Tables A3 and A4), showed measurable benefit from RMM. While RFNet achieves competitive HD95 scores without augmentation, training with RMM consistently enhanced spatial boundary delineation, particularly in ET and TC, suggesting complementary effects rather than redundancy with strong architectural priors.

Figure 3 presents qualitative comparisons under missing modality settings, where M^2^FTrans trained with RMM generated more anatomically consistent segmentations. DSC scores for the three tumor regions were included within the coronal slices corresponding to each prediction, while the sagittal views displayed DSC scores for the NCR/NET and ED subregions. RMM consistently produced higher DSC scores for NCR/NET, a class known for subtle intensity differences and structural complexity, demonstrating its effectiveness in learning fine-grained anatomical priors.

Finally, we compared models trained with RMM to state-of-the-art methods trained on larger regions of interest (ROI=80) in Supplementary Table A5. Despite using a smaller ROI (64), our method achieved competitive performance with the state-of-the-art across all subregions. These findings are particularly relevant in clinical settings where computational and time constraints limit the size of processable image volumes, reinforcing the value of our proposed framework in resource-constrained environments.

## 6 Discussion

This study presents a flexible image segmentation framework that robustly handles missing MRI sequences during training and inference. Central to our framework is *Region ModalMix* (RMM), a novel augmentation strategy that leverages brain parcellation masks to mix information across modalities within anatomical regions. In so doing, RMM enhances the model’s ability to learn spatially coherent, modality-agnostic features that generalize across a wide range of missing data scenarios.

Our segmentation results demonstrate that RMM consistently outperforms existing augmentation methods such as 3D MM-CutMix [27] across different metrics, especially in the enhancing tumor and necrotic/non-enhancing tumor regions. These regions are clinically important but challenging to delineate due to their small size and heterogeneous appearance [13]. Notably, RMM achieved marked gains in boundary precision, indicating better anatomical fidelity, which is a key attribute for real-world deployment where segmentation errors at the margins can influence treatment decisions. Furthermore, RMM improved segmentation performance without requiring increased input volume size, making it computationally efficient and suitable for deployment in environments with limited resources. Our approach is also agnostic to architectural choices, with significant performance boosts across diverse backbones, including transformer-based (e.g., mmFormer, IMS^2^Trans) and hybrid models.

Our framework has some limitations. First, RMM currently relies on pre-computed anatomical parcellations, which may not be equally accurate across populations (e.g., pediatric or atypical brain anatomies). Second, while we evaluated our framework across multiple backbones, we focused on the task of brain tumor segmentation from a limited set of MRI sequences (i.e., T1, T1ce, T2, FLAIR), but other modalities such as diffusion-weighted imaging and susceptibility-weighted imaging, were not explored due to data availability constraints. Including these additional modalities in future work would provide a more comprehensive assessment of the framework’s scalability and utility. Third, the augmentations are static and do not adapt to task-specific features or pathologies. Finally, while we simulate missing modalities, further validation on large-scale prospective datasets with naturally missing data is needed to confirm external validity.

In conclusion, our proposed anatomically guided, modality-agnostic augmentation strategy is designed to improve robustness and generalizability in multimodal brain MRI analysis. By structurally mixing anatomical regions across modalities, our approach enables deep learning models to maintain high performance even when key MRI sequences are missing. Through comprehensive evaluation on brain tumor segmentation, we demonstrated that our method improves segmentation accuracy and predictive reliability across diverse input configurations and model backbones. Our framework also demonstrated better volumetric overlap and boundary precision on smaller structures. These findings highlight the promise of anatomy-aware training as a foundation for developing clinically resilient neuroimaging AI systems capable of working effectively in real-world, heterogeneous data environments.

## Data Availability

https://www.med.upenn.edu/cbica/brats2020/data.html

## Author contributions

DL, DA, and VBK: study conception and design. DL and DA: data collection and processing. DL and DA: implementation. DL and DA: analysis. DL, DA, SAB, BAP, and VBK: data interpretation and manuscript write-up. VBK: study direction. All authors reviewed the results and approved the final version of the manuscript.

## Funding

This project was supported by grants from the National Institute on Aging’s Artificial Intelligence and Technology Collaboratories (P30-AG073104 and P30-AG073105), and the National Institutes of Health (R01-HL159620, R01-AG083735, R01-AG062109, and R01-NS142076).

## Notes

### Competing Interest Statement

V.B.K. is a co-founder and equity holder of deepPath Inc., and CogniScreen, Inc. He also serves on the scientific advisory board of Altoida Inc. The remaining authors declare no competing interests.

### Funding Statement

This project was supported by grants from the National Institute on Aging Artificial Intelligence and Technology Collaboratories (P30-AG073104 and P30-AG073105), and the National Institutes of Health (R01-HL159620,
R01-AG083735, R01-AG062109, and R01-NS142076).

### Author Declarations

https://www.med.upenn.edu/cbica/brats2020/data.html

