## Supplement for "Anatomy-guided, modality-agnostic segmentation of neuroimaging abnormalities"

### A Brain parcellation labels

To obtain consistent anatomical segmentations across MRI scans, we utilized **SynthSeg+** [1], a domain-agnostic deep learning based segmentation framework for brain MRI. The SynthSeg+ framework employs a hierarchical, modular CNN architecture designed for robust whole-brain segmentation of heterogeneous clinical brain MRI scans. The architecture is composed of four main modules: a coarse tissue segmenter ($S_1$), a denoiser ($D$), a fine-grained structure segmenter ($S_2$), and a cortical parcellation module ($S_3$). The first network, $S_1$, segments the input scan into four coarse tissue classes (cortical gray matter, white matter, CSF, and cerebellum). These preliminary labels are passed through a denoising network $D$, which corrects topological inconsistencies and segmentation errors introduced by low-quality input scans. The refined output is then fed, along with the original image, into a second segmentation network $S_2$, which predicts high-resolution segmentations of 31 anatomical regions. For further granularity, especially in the cortex, a third segmenter $S_3$ parcellates the bilateral cortical masks produced by $S_2$ into 68 cortical subregions. Additionally, a regression module ($R$) estimates region-specific segmentation quality (Dice proxy scores), enabling automated quality control (QC). All networks are implemented as modified 3D U-Nets with Exponential Linear Unit (ELU) activations and are trained independently using synthetic data generated via domain randomization, ensuring robustness to arbitrary MR contrast, resolution, orientation, and noise levels. Table A1 includes a list of all the parcellated brain structures ($n = 101$) that were utilized by our proposed Region ModalMix as anatomical priors.

### B Experimental setup

Model settings for the brain tumor segmentation task are detailed in Table A2. Models were trained with preserved latent dimensionality as introduced in [2, 5, 3, 4]. The difference in trainable parameter count between our trained models and those reported in the literature is attributed to the smaller region-of-interest size of 64 that we adopted for training.

### C Additional results

State-of-the-art brain tumor segmentation methods typically report performance on the three standard composite tumor regions—whole tumor (WT), tumor core (TC), and enhancing tumor (ET)—while often omitting evaluation on finer-grained subregions. To complement the main results and provide a more granular performance assessment, we conducted an in-depth analysis of our

Table A1: **Anatomical label definitions used by SynthSeg+.** The anatomical brain region labels are reported along with their associated numerical identifiers as defined by SynthSeg+ [1]. Superscripts $^{L/R}$ indicate the left and right hemispheres, respectively. The labels encompass a comprehensive set of cortical and subcortical structures, ventricles, and white matter regions across both hemispheres, supporting detailed whole-brain segmentation.

| Label | Value | Label | Value |
|---|---|---|---|
| background | 0 | cerebral white matter$^{L/R}$ | 2/41 |
| cerebral cortex$^{L/R}$ | 3/42 | lateral ventricle$^{L/R}$ | 4/43 |
| inferior lateral ventricle$^{L/R}$ | 5/44 | cerebellum white matter$^{L/R}$ | 7/46 |
| cerebellum cortex$^{L/R}$ | 8/47 | thalamus$^{L/R}$ | 10/49 |
| caudate$^{L/R}$ | 11/50 | putamen$^{L/R}$ | 12/51 |
| pallidum$^{L/R}$ | 13/52 | 3rd ventricle | 14 |
| 4th ventricle | 15 | brain-stem | 16 |
| hippocampus$^{L/R}$ | 17/53 | amygdala$^{L/R}$ | 18/54 |
| accumbens area$^{L/R}$ | 26/58 | CSF | 24 |
| ventral DC$^{L/R}$ | 28/60 | banks of the superior temporal sulcus$^{L/R}$ | 1001/2001 |
| caudal anterior cingulate cortex$^{L/R}$ | 1002/2002 | caudal middle frontal gyrus$^{L/R}$ | 1003/2003 |
| cuneus cortex$^{L/R}$ | 1005/2005 | entorhinal cortex$^{L/R}$ | 1006/2006 |
| fusiform gyrus$^{L/R}$ | 1007/2007 | inferior parietal lobule$^{L/R}$ | 1008/2008 |
| inferior temporal gyrus$^{L/R}$ | 1009/2009 | isthmus of the cingulate cortex$^{L/R}$ | 1010/2010 |
| lateral occipital cortex$^{L/R}$ | 1011/2011 | lateral orbitofrontal cortex$^{L/R}$ | 1012/2012 |
| lingual gyrus$^{L/R}$ | 1013/2013 | medial orbitofrontal cortex$^{L/R}$ | 1014/2014 |
| middle temporal gyrus$^{L/R}$ | 1015/2015 | parahippocampal gyrus$^{L/R}$ | 1016/2016 |
| paracentral lobule$^{L/R}$ | 1017/2017 | pars opercularis$^{L/R}$ | 1018/2018 |
| pars orbitalis$^{L/R}$ | 1019/2019 | pars triangularis$^{L/R}$ | 1020/2020 |
| pericalcarine cortex$^{L/R}$ | 1021/2021 | postcentral gyrus$^{L/R}$ | 1022/2022 |
| posterior cingulate cortex$^{L/R}$ | 1023/2023 | precentral gyrus$^{L/R}$ | 1024/2024 |
| precuneus cortex$^{L/R}$ | 1025/2025 | rostral anterior cingulate cortex$^{L/R}$ | 1026/2026 |
| rostral middle frontal gyrus$^{L/R}$ | 1027/2027 | superior frontal gyrus$^{L/R}$ | 1028/2028 |
| superior parietal lobule$^{L/R}$ | 1029/2029 | superior temporal gyrus$^{L/R}$ | 1030/2030 |
| supramarginal gyrus$^{L/R}$ | 1031/2031 | frontal pole$^{L/R}$ | 1032/2032 |
| temporal pole$^{L/R}$ | 1033/2033 | transverse temporal gyrus$^{L/R}$ | 1034/2034 |
| insular cortex$^{L/R}$ | 1035/2035 | | |

Table A2: **Model settings used for training on brain tumor segmentation.** The number of trainable parameters and the latent dimensionality were reported for each of the model architectures included in our study.

| **Architecture** | **Trainable parameters** | **Latent dimension** |
|---|---|---|
| RFNet [2] | 8.9M | 128 |
| mmFormer [5] | 35.7M | 512 |
| IMS$^2$Trans [4] | 3.6M | 192 |
| M$^2$FTrans [3] | 45.3M | 512 |

framework's performance effectiveness across all relevant tumor compartments, including edema (ED) and necrotic/non-enhancing tumor (NCR/NET).

Tables A3 and A4 present the 95$^{\text{th}}$ percentile Hausdorff Distance (HD95) and the Dice Similarity Coefficient (DSC) scores, respectively, across all evaluated models and MRI modality combinations. Across both metrics, models augmented with our proposed Region ModalMix (RMM) consistently outperformed their baseline counterparts, demonstrating the augmentation method's robustness and generalizability. Performance gains were observed across multiple architectures, including RFNet [2], mmFormer [5], $M^2$FTrans [3], and IMS$^2$Trans [4], underscoring the broad applicability of our approach.

To ensure a fair and comprehensive comparison with prior work, we also reported results on two region-of-interest (ROI) (64 and 80 voxels) in Table A5. While our primary experiments were conducted using ROI=64 to reduce computational requirements and enable faster training, previous methods commonly adopted a larger ROI of 80 [2, 5, 3]. To bridge this gap, we reproduced results for mmFormer and $M^2$FTrans using ROI=80. Notably, $M^2$FTrans achieved the strongest performance in this setting, particularly in terms of average DSC and HD95 across all tumor regions. These findings indicate that even under a more resource-efficient configuration, our framework delivers performance almost on par with that of larger-scale baselines, offering a favorable trade-off between computational efficiency and segmentation quality. This is particularly relevant in real-world settings where computational and time constraints limit the size of processable image volumes.

Table A3: **Performance comparison across 15 different MRI modality combinations of the BraTS 2020 dataset, evaluated by the 95th percentile Hausdorff Distance (HD95, mm).** Lower values indicate better performance. Each row corresponds to a model trained either with or without our proposed augmentation strategy. The '3CM' suffix refers to models enhanced with 3D MM-CutMix. For each combination, the best result was shown in **bold** and the second best was underlined. All comparisons were stratified by tumor subregion: whole tumor (WT), tumor core (TC), enhancing tumor (ET), non-enhancing tumor (NCR/NET), and edema (ED).

| Type | FLAIR | ○ | ○ | ○ | ● | ○ | ○ | ● | ○ | ● | ● | ● | ● | ● | ○ | ● | Avg. | p-value |
|---|---|---|---|---|---|---|---|---|---|---|---|---|---|---|---|---|---|---|
| | T1 | ○ | ○ | ● | ○ | ○ | ● | ● | ● | ○ | ○ | ● | ● | ○ | ● | ● | | |
| | T1ce | ○ | ● | ○ | ○ | ● | ● | ○ | ○ | ○ | ● | ● | ○ | ● | ● | ● | | |
| | T2 | ● | ○ | ○ | ○ | ● | ○ | ○ | ● | ● | ○ | ○ | ● | ● | ● | ● | | |
| WT | RFNet [2] | 27.55 | 14.07 | 15.85 | 17.42 | 12.61 | 9.53 | 9.52 | 8.16 | 13.89 | 9.27 | 6.42 | 7.61 | 9.36 | 6.71 | 5.60 | 11.57 | < 0.001 |
| | RFNet [2] w/ Ours | 16.43 | 14.01 | 12.97 | 20.29 | 8.15 | 10.93 | 7.26 | 8.15 | 9.84 | 9.18 | 5.95 | 7.33 | 8.22 | 6.53 | 6.71 | 10.13 | < 0.001 |
| | mmFormer [5] | 31.82 | 15.50 | 17.03 | 34.40 | 22.44 | 10.30 | 11.75 | 11.98 | 28.55 | 21.57 | 8.22 | 12.20 | 20.14 | 7.47 | 7.99 | 17.42 | < 0.001 |
| | mmFormer [5] w/ Ours | 20.31 | **11.07** | 21.27 | 15.87 | 14.86 | 10.52 | 7.39 | 9.40 | 8.65 | 11.19 | 6.78 | 5.88 | 7.75 | 5.95 | 5.45 | 10.82 | < 0.001 |
| | M$^2$FTrans [3] | 12.34 | 12.75 | 27.63 | 40.30 | 18.17 | 10.61 | 6.65 | 6.91 | 25.82 | 31.26 | 15.26 | 6.89 | 26.94 | 8.76 | 15.47 | 17.72 | < 0.001 |
| | M$^2$FTrans [3] w/ Ours | 29.96 | 12.18 | **12.64** | 17.05 | 20.42 | **8.79** | 7.14 | 8.22 | 18.04 | 7.65 | 5.59 | 7.27 | 9.87 | 8.05 | 5.23 | 11.87 | < 0.001 |
| | IMS$^2$Trans [4] | 8.55 | 21.80 | 15.46 | 9.55 | 7.74 | 12.80 | **5.76** | 6.30 | **5.11** | 7.45 | **5.57** | **4.73** | **5.10** | 7.37 | **4.46** | 8.52 | < 0.001 |
| | IMS$^2$Trans [4] w/ 3CM | 8.11 | 19.09 | 14.60 | 9.28 | **5.80** | 14.15 | 6.32 | 6.18 | 5.60 | 6.18 | 5.78 | 5.21 | 5.52 | 5.56 | 5.37 | 8.18 | < 0.001 |
| | IMS$^2$Trans [4] w/ Ours | **6.64** | 17.73 | 14.83 | **9.12** | 7.93 | 14.65 | 6.58 | **5.38** | 6.46 | **6.07** | **5.57** | 5.45 | 5.66 | **5.43** | 4.49 | **8.13** | < 0.001 |
| TC | RFNet [2] | 12.15 | 11.39 | 20.89 | 16.65 | 17.82 | **5.68** | **8.33** | 8.36 | 9.32 | 14.89 | **7.01** | 8.21 | 15.98 | 6.80 | 6.47 | 11.33 | < 0.001 |
| | RFNet [2] w/ Ours | **8.88** | 12.12 | 15.06 | **13.56** | 8.36 | 6.73 | 8.78 | 7.61 | **9.10** | 8.70 | 7.24 | 8.49 | 8.53 | **4.95** | 6.25 | **8.96** | < 0.001 |
| | mmFormer [5] | 23.42 | 12.47 | 27.60 | 39.26 | 36.45 | 6.78 | 12.37 | 11.27 | 27.10 | 36.12 | 9.93 | 11.93 | 36.77 | 7.43 | 12.38 | 20.75 | < 0.001 |
| | mmFormer [5] w/ Ours | 18.58 | 13.17 | 31.84 | 21.12 | 27.48 | 8.65 | 9.25 | 13.66 | 11.91 | 17.92 | 7.33 | 8.62 | 15.90 | 8.30 | 7.07 | 14.72 | < 0.001 |
| | M$^2$FTrans [3] | 11.58 | **11.06** | 35.32 | 41.10 | 30.48 | 7.66 | 9.77 | **7.46** | 15.28 | 48.05 | 26.37 | 8.45 | 44.20 | 13.75 | 26.73 | 22.48 | < 0.001 |
| | M$^2$FTrans [3] w/ Ours | 16.44 | 12.23 | 15.78 | 15.91 | 19.96 | 5.70 | 10.04 | 9.67 | 12.08 | 12.86 | 5.55 | **7.52** | 13.35 | 6.92 | **5.94** | 11.33 | < 0.001 |
| | IMS$^2$Trans [4] | 11.23 | 25.12 | 13.59 | 15.68 | 6.33 | 9.67 | 11.13 | 9.31 | 10.40 | 9.84 | 7.41 | 9.16 | **6.58** | 6.97 | 6.44 | 10.59 | < 0.001 |
| | IMS$^2$Trans [4] w/ 3CM | 10.99 | 19.97 | **13.01** | 13.88 | **5.62** | 8.13 | 11.01 | 9.25 | 10.77 | 9.21 | 7.39 | 11.59 | 8.03 | 5.73 | 6.70 | 10.09 | < 0.001 |
| | IMS$^2$Trans [4] w/ Ours | 11.29 | 18.43 | 13.07 | 14.32 | 7.16 | 9.47 | 10.17 | 8.76 | 10.60 | **8.26** | 7.45 | 8.87 | 8.05 | 5.26 | 6.48 | 9.84 | < 0.001 |
| ET | RFNet [2] | 8.44 | 9.03 | 11.63 | 10.74 | 12.84 | 2.36 | 7.18 | 5.82 | 6.78 | 8.50 | 4.45 | 5.20 | 12.06 | 2.71 | 3.83 | 7.44 | < 0.001 |
| | RFNet [2] w/ Ours | **6.30** | 8.84 | 9.55 | 10.21 | 4.20 | 2.92 | 8.34 | **5.69** | **5.97** | **5.23** | 3.81 | **5.14** | 4.61 | **1.96** | 4.72 | **5.83** | < 0.001 |
| | mmFormer [5] | 12.08 | 9.33 | 19.45 | 22.38 | 30.48 | 2.54 | 8.87 | 8.06 | 11.05 | 27.86 | 6.08 | 8.30 | 30.23 | 2.91 | 8.01 | 13.84 | < 0.001 |
| | mmFormer [5] w/ Ours | 9.61 | 9.61 | 24.62 | 8.89 | 22.11 | 3.41 | 6.15 | 8.57 | 7.53 | 13.99 | 2.54 | 5.37 | 10.31 | 5.07 | 3.59 | 9.42 | < 0.001 |
| | M$^2$FTrans [3] | 9.05 | 9.03 | 27.57 | 21.02 | 24.13 | 3.86 | 7.71 | 5.93 | 9.45 | 40.00 | 22.31 | 7.07 | 33.94 | 10.34 | 22.45 | 16.92 | < 0.001 |
| | M$^2$FTrans [3] w/ Ours | 9.47 | **8.52** | 9.42 | **7.59** | 16.52 | **2.16** | **6.10** | 7.06 | 8.35 | 7.30 | **2.20** | 5.21 | 9.36 | 3.28 | **2.22** | 6.98 | < 0.001 |
| | IMS$^2$Trans [4] | 7.28 | 19.12 | **5.88** | 9.45 | 3.63 | 5.64 | 8.44 | 7.14 | 6.73 | 6.54 | 4.75 | 6.45 | **3.61** | 3.01 | 3.64 | 6.75 | < 0.001 |
| | IMS$^2$Trans [4] w/ 3CM | 7.36 | 15.48 | 9.01 | 9.24 | **3.27** | 5.45 | 7.51 | 6.87 | 8.32 | 7.31 | 3.86 | 8.28 | 5.48 | 2.72 | 3.80 | 6.93 | < 0.001 |
| | IMS$^2$Trans [4] w/ Ours | 7.72 | 12.16 | 7.67 | 9.87 | 3.77 | 4.85 | 8.05 | 5.80 | 8.49 | 6.00 | 3.11 | 7.79 | 4.86 | 1.97 | 3.99 | 6.40 | < 0.001 |
| NCR/NET | RFNet [2] | 13.47 | 10.17 | 11.13 | 17.49 | 8.10 | 7.01 | **8.75** | **8.28** | **9.65** | 8.43 | **6.31** | 8.33 | 7.63 | 6.93 | **6.14** | 9.19 | < 0.001 |
| | RFNet [2] w/ Ours | 10.25 | **9.70** | **8.49** | **11.66** | 7.25 | 7.34 | 9.38 | 8.83 | 10.06 | 7.65 | 7.41 | 8.32 | **7.53** | **6.52** | 6.57 | **8.46** | < 0.001 |
| | mmFormer [5] | 23.45 | 10.53 | 12.88 | 39.28 | 15.51 | 7.27 | 12.95 | 9.42 | 29.07 | 16.20 | 8.81 | 11.35 | 21.53 | 8.62 | 9.88 | 15.78 | < 0.001 |
| | mmFormer [5] w/ Ours | 17.34 | 13.64 | 11.73 | 21.40 | 10.33 | 7.77 | 10.02 | 9.96 | 12.22 | 10.62 | 8.02 | 9.98 | 10.39 | 7.63 | 7.20 | 11.22 | < 0.001 |
| | M$^2$FTrans [3] | **9.97** | 11.47 | 18.64 | 38.10 | 17.92 | **6.67** | 9.82 | 8.86 | 13.38 | 33.31 | 14.33 | **8.25** | 31.74 | 9.13 | 16.90 | 16.56 | < 0.001 |
| | M$^2$FTrans [3] w/ Ours | 17.64 | 10.93 | 8.89 | 16.47 | 10.59 | 6.97 | 11.09 | 10.74 | 11.92 | **6.86** | 6.65 | 8.70 | 8.85 | 7.51 | 7.19 | 10.07 | < 0.001 |
| | IMS$^2$Trans [4] | 11.09 | 18.47 | 14.06 | 16.10 | **7.21** | 10.52 | 10.18 | 9.59 | 10.98 | 12.48 | 9.01 | 9.37 | 9.26 | 6.98 | 7.37 | 10.84 | < 0.001 |
| | IMS$^2$Trans [4] w/ 3CM | 11.00 | 18.23 | 11.16 | 14.21 | 7.27 | 8.32 | 10.23 | 9.28 | 10.66 | 9.92 | 9.75 | 11.17 | 9.00 | 7.29 | 8.12 | 10.37 | < 0.001 |
| | IMS$^2$Trans [4] w/ Ours | 10.72 | 17.70 | 12.18 | 14.47 | 8.16 | 9.39 | 10.58 | 9.27 | 10.62 | 8.92 | 8.02 | 9.40 | 7.93 | 6.78 | 7.78 | 10.13 | < 0.001 |
| ED | RFNet [2] | 26.29 | 14.06 | 11.20 | 17.13 | 8.00 | 9.60 | 10.15 | 8.88 | 12.99 | 9.01 | 5.64 | 8.15 | 7.02 | 5.99 | 5.52 | 10.64 | < 0.001 |
| | RFNet [2] w/ Ours | 15.82 | 13.96 | 10.86 | 19.09 | 7.26 | 11.01 | 7.94 | 8.52 | 10.47 | 8.97 | 6.65 | 8.60 | 7.17 | 6.93 | 6.73 | 10.00 | < 0.001 |
| | mmFormer [5] | 27.24 | 15.00 | 11.06 | 29.90 | 9.64 | 9.83 | 11.85 | 11.64 | 24.87 | 13.23 | 7.64 | 12.06 | 9.65 | 7.58 | 6.72 | 13.86 | < 0.001 |
| | mmFormer [5] w/ Ours | 18.64 | **11.08** | 10.86 | 14.47 | 6.17 | 8.59 | 8.34 | 9.08 | 8.64 | 7.88 | 7.04 | 6.63 | 5.17 | 5.80 | 4.98 | 8.89 | < 0.001 |
| | M$^2$FTrans [3] | 11.90 | 12.60 | 13.87 | 36.87 | 8.87 | 10.38 | 6.61 | 7.30 | 23.00 | 15.98 | 7.82 | 7.80 | 11.62 | 6.52 | 8.41 | 12.64 | < 0.001 |
| | M$^2$FTrans [3] w/ Ours | 28.79 | 12.14 | **10.03** | 15.53 | 16.83 | **8.13** | 7.91 | 8.50 | 17.10 | 6.77 | 5.44 | 7.43 | 7.92 | 7.27 | 5.34 | 11.01 | < 0.001 |
| | IMS$^2$Trans [4] | 8.52 | 20.22 | 14.93 | 10.44 | 6.98 | 11.18 | 6.69 | 7.27 | **6.46** | 6.75 | 5.72 | 6.05 | 5.44 | 7.54 | 4.77 | 8.60 | < 0.001 |
| | IMS$^2$Trans [4] w/ 3CM | 8.03 | 18.49 | 13.97 | **9.58** | **5.89** | 12.58 | 7.26 | 7.12 | 6.74 | 6.22 | 5.92 | 6.41 | 5.69 | 5.80 | 5.20 | 8.33 | < 0.001 |
| | IMS$^2$Trans [4] w/ Ours | **7.27** | 17.30 | 14.12 | 9.83 | 7.27 | 12.94 | **6.60** | **6.47** | 7.12 | **5.73** | **5.23** | **5.99** | **4.70** | **5.77** | **4.52** | **8.06** | < 0.001 |

Table A4: **Performance comparison across 15 different MRI modality combinations of the BraTS 2020 dataset, evaluated by the Dice Similarity Coefficient score (DSC, %).** Higher values indicate better performance. Each row corresponds to a model trained either with or without our proposed augmentation strategy. The '3CM' suffix refers to models enhanced with 3D MM-CutMix. For each combination, the best result was shown in **bold** and the second best was underlined. All comparisons were stratified by tumor subregion: whole tumor (WT), tumor core (TC), enhancing tumor (ET), non-enhancing tumor (NCR/NET), and edema (ED).

| Type | | | | | | | | | | | | | | | | | Avg. | p-value |
|---|---|---|---|---|---|---|---|---|---|---|---|---|---|---|---|---|---|---|
| | FLAIR | ○ | ○ | ○ | ● | ○ | ○ | ● | ○ | ● | ● | ● | ● | ● | ○ | ● | | |
| | T1 | ○ | ○ | ● | ○ | ○ | ● | ● | ● | ○ | ○ | ● | ● | ○ | ● | ● | | |
| | T1ce | ○ | ● | ○ | ○ | ● | ● | ○ | ○ | ○ | ● | ● | ○ | ● | ● | ● | | |
| | T2 | ● | ○ | ○ | ○ | ● | ○ | ○ | ● | ● | ○ | ○ | ● | ● | ● | ● | | |
| WT | RFNet [2] | 83.70 | **75.62** | 75.72 | 84.85 | **87.27** | **80.01** | 89.15 | **87.69** | **88.81** | **89.16** | **90.22** | **90.29** | 89.89 | **88.34** | **91.01** | **86.12** | < 0.001 |
| | RFNet [2] w/ Ours | 83.28 | 72.34 | 75.28 | 84.26 | 86.72 | 78.82 | 88.41 | 86.01 | 88.32 | 88.52 | 89.24 | 89.57 | 89.66 | 87.44 | 90.08 | 85.20 | < 0.001 |
| | mmFormer [5] | 80.50 | 72.78 | 73.93 | 79.93 | 84.65 | 78.19 | 86.67 | 85.48 | 84.66 | 86.01 | 88.47 | 87.93 | 87.08 | 86.80 | 89.13 | 83.48 | < 0.001 |
| | mmFormer [5] w/ Ours | 82.91 | 72.82 | 72.74 | 84.11 | 85.54 | 77.67 | 87.49 | 86.09 | 87.58 | 87.85 | 88.68 | 88.84 | 88.86 | 87.04 | 89.46 | 84.51 | < 0.001 |
| | M$^2$FTrans [3] | **85.12** | 74.25 | 74.77 | 79.91 | 85.97 | 79.69 | 88.74 | 86.67 | 86.74 | 84.98 | 88.26 | 89.71 | 87.13 | 87.35 | 88.84 | 84.54 | < 0.001 |
| | M$^2$FTrans [3] w/ Ours | 81.21 | 74.20 | **75.75** | 84.66 | 85.94 | 79.84 | **89.43** | 86.93 | 87.77 | 88.97 | 89.99 | 90.16 | 89.48 | 88.05 | 90.58 | 85.53 | < 0.001 |
| | IMS$^2$Trans [4] | 84.28 | 69.13 | 72.45 | 84.75 | 87.03 | 75.93 | 88.14 | 86.78 | 88.72 | 88.37 | 89.04 | 89.93 | **90.00** | 87.45 | 90.46 | 84.83 | < 0.001 |
| | IMS$^2$Trans [4] w/ 3CM | 83.68 | 68.40 | 73.52 | **85.62** | 86.60 | 76.04 | 88.11 | 86.12 | 88.21 | 88.23 | 88.71 | 89.21 | 89.32 | 87.04 | 89.62 | 84.56 | < 0.001 |
| | IMS$^2$Trans [4] w/ Ours | 83.87 | 68.28 | 71.99 | 84.42 | 86.66 | 75.81 | 88.43 | 86.32 | 88.69 | 88.79 | 89.61 | 89.79 | 89.89 | 87.55 | 90.44 | 84.70 | < 0.001 |
| TC | RFNet [2] | 68.99 | **65.81** | 78.77 | **67.29** | 81.74 | 82.58 | **73.26** | 72.56 | 73.29 | 83.17 | 84.33 | 75.19 | 83.83 | 84.09 | 85.12 | **77.34** | < 0.001 |
| | RFNet [2] w/ Ours | 66.97 | 56.10 | **80.04** | 65.22 | **83.58** | 81.64 | 69.71 | 69.57 | 71.33 | **84.06** | 84.86 | 72.64 | **84.67** | 83.99 | **85.18** | 75.97 | < 0.001 |
| | mmFormer [5] | 66.44 | 62.80 | 76.97 | 61.64 | 77.70 | 81.82 | 71.15 | 70.58 | 69.09 | 77.55 | 82.44 | 72.56 | 78.22 | 82.60 | 82.13 | 74.25 | < 0.001 |
| | mmFormer [5] w/ Ours | 64.71 | 58.60 | 74.40 | 60.96 | 79.30 | 80.73 | 69.47 | 69.97 | 69.02 | 82.71 | 84.12 | 72.33 | 83.36 | 82.92 | 84.29 | 74.46 | < 0.001 |
| | M$^2$FTrans [3] | **71.47** | 64.40 | 76.25 | 63.56 | 79.11 | 82.87 | 72.75 | **73.25** | **73.53** | 74.94 | 80.88 | **75.29** | 77.09 | 82.99 | 80.53 | 75.26 | < 0.001 |
| | M$^2$FTrans [3] w/ Ours | 66.61 | 62.75 | 79.47 | 65.59 | 81.51 | **82.95** | 73.07 | 71.17 | 72.22 | 83.69 | **85.42** | 75.13 | 83.21 | **84.49** | 85.11 | 76.83 | < 0.001 |
| | IMS$^2$Trans [4] | 62.90 | 54.75 | 76.06 | 63.37 | 82.25 | 79.94 | 68.52 | 67.65 | 68.31 | 81.60 | 82.87 | 70.04 | 82.90 | 83.14 | 83.64 | 73.86 | < 0.001 |
| | IMS$^2$Trans [4] w/ 3CM | 63.71 | 52.34 | 77.59 | 63.82 | 81.69 | 80.26 | 68.57 | 67.30 | 68.73 | 82.31 | 82.86 | 70.17 | 82.75 | 81.92 | 82.78 | 73.79 | < 0.001 |
| | IMS$^2$Trans [4] w/ Ours | 59.52 | 47.13 | 75.10 | 58.23 | 81.71 | 80.31 | 67.09 | 65.78 | 66.50 | 82.41 | 83.57 | 70.22 | 82.61 | 82.81 | 82.97 | 72.40 | < 0.001 |
| ET | RFNet [2] | 46.04 | 34.68 | 73.99 | 38.64 | 74.74 | **80.26** | 42.44 | 45.78 | 47.99 | 76.48 | 77.96 | 50.54 | **76.38** | 78.68 | 78.38 | 61.53 | < 0.001 |
| | RFNet [2] w/ Ours | **48.02** | 36.30 | **74.93** | 38.68 | 76.41 | 78.89 | 44.99 | 50.71 | 48.76 | 74.86 | 76.93 | **52.51** | 75.73 | 78.90 | 79.38 | **62.40** | < 0.001 |
| | mmFormer [5] | 46.00 | 33.72 | 71.72 | 34.15 | 67.51 | 76.64 | 40.38 | 46.22 | 48.02 | 68.87 | 75.54 | 49.03 | 68.32 | 76.10 | 74.05 | 58.42 | < 0.001 |
| | mmFormer [5] w/ Ours | 46.26 | 33.01 | 67.21 | 30.95 | 69.91 | 76.44 | 39.93 | 47.40 | 46.68 | 71.76 | 79.73 | 48.15 | 73.23 | 77.55 | **80.29** | 59.23 | < 0.001 |
| | M$^2$FTrans [3] | 47.48 | **38.56** | 68.11 | **38.90** | 72.16 | 77.54 | **47.16** | **50.89** | 50.57 | 62.68 | 71.96 | 51.54 | 65.49 | 77.86 | 71.06 | 59.46 | < 0.001 |
| | M$^2$FTrans [3] w/ Ours | 46.97 | 34.11 | 71.75 | 36.70 | 74.17 | 78.31 | 43.41 | 48.28 | **51.16** | 73.40 | 78.97 | 50.89 | 75.51 | **79.60** | 79.94 | 61.54 | < 0.001 |
| | IMS$^2$Trans [4] | 38.42 | 25.91 | 70.90 | 33.80 | 76.15 | 74.77 | 37.34 | 41.17 | 40.39 | 74.37 | 75.82 | 42.35 | 75.08 | 76.38 | 76.11 | 57.26 | < 0.001 |
| | IMS$^2$Trans [4] w/ 3CM | 39.81 | 23.99 | 72.76 | 32.45 | 74.86 | 71.93 | 35.74 | 40.74 | 39.44 | 73.81 | 75.76 | 42.84 | 74.91 | 74.56 | 75.79 | 56.63 | < 0.001 |
| | IMS$^2$Trans [4] w/ Ours | 39.76 | 23.22 | 70.79 | 31.16 | **77.02** | 75.86 | 36.84 | 42.58 | 43.34 | **77.64** | **79.78** | 45.67 | 74.61 | 78.91 | 78.69 | 58.39 | < 0.001 |
| NCR/NET | RFNet [2] | 47.75 | **46.59** | **64.37** | **41.56** | **66.49** | 67.05 | **51.73** | **51.30** | **51.86** | 66.69 | 67.53 | **53.99** | **68.02** | **68.09** | **68.42** | **58.76** | < 0.001 |
| | RFNet [2] w/ Ours | 44.17 | 42.31 | 62.68 | 41.09 | 65.75 | 64.18 | 49.40 | 48.73 | 48.34 | **67.01** | 67.53 | 50.43 | 67.10 | 65.76 | 67.34 | 56.79 | < 0.001 |
| | mmFormer [5] | 44.93 | 43.69 | 62.25 | 38.17 | 63.99 | 66.00 | 48.86 | 49.75 | 46.88 | 63.74 | 65.67 | 50.82 | 64.02 | 65.34 | 65.60 | 55.98 | < 0.001 |
| | mmFormer [5] w/ Ours | 42.57 | 41.05 | 62.50 | 39.31 | 64.61 | 64.76 | 48.08 | 49.79 | 48.75 | 65.87 | **68.14** | 51.25 | 66.13 | 66.59 | 67.47 | 56.46 | < 0.001 |
| | M$^2$FTrans [3] | **48.05** | 44.48 | 64.07 | 37.54 | 64.37 | **67.27** | 50.12 | 50.35 | 49.22 | 62.75 | 66.24 | 51.52 | 63.52 | 66.26 | 65.65 | 56.76 | < 0.001 |
| | M$^2$FTrans [3] w/ Ours | 44.30 | 45.24 | 64.30 | 41.09 | 65.52 | 66.38 | 50.15 | 50.18 | 50.87 | 66.85 | 67.78 | 52.60 | 66.57 | 66.66 | 67.56 | 57.74 | < 0.001 |
| | IMS$^2$Trans [4] | 44.38 | 40.99 | 59.90 | 37.90 | 64.70 | 63.83 | 47.54 | 48.91 | 47.35 | 63.69 | 64.81 | 49.45 | 65.12 | 65.57 | 65.73 | 55.32 | < 0.001 |
| | IMS$^2$Trans [4] w/ 3CM | 44.10 | 39.50 | 61.68 | 39.30 | 63.74 | 64.61 | 47.92 | 48.96 | 48.26 | 64.62 | 65.77 | 49.75 | 65.04 | 64.50 | 65.23 | 55.53 | < 0.001 |
| | IMS$^2$Trans [4] w/ Ours | 42.11 | 36.39 | 58.91 | 34.75 | 64.11 | 63.78 | 47.18 | 46.02 | 45.40 | 65.19 | 66.69 | 48.84 | 65.41 | 65.25 | 65.79 | 54.39 | < 0.001 |
| ED | RFNet [2] | 63.04 | **54.73** | 58.13 | **68.04** | 72.56 | **63.05** | 71.69 | 68.87 | **71.61** | 76.32 | 77.21 | 73.68 | **77.33** | 73.64 | **78.41** | **69.89** | < 0.001 |
| | RFNet [2] w/ Ours | 63.50 | 52.20 | 56.68 | 67.13 | 71.59 | 60.76 | 70.77 | 68.38 | 70.48 | 75.67 | 76.43 | 72.40 | 76.85 | 72.61 | 77.69 | 68.88 | < 0.001 |
| | mmFormer [5] | 60.33 | 51.02 | 56.52 | 63.45 | 71.03 | 60.96 | 68.44 | 66.66 | 67.43 | 74.10 | 75.44 | 70.31 | 75.43 | 72.17 | 76.11 | 67.29 | < 0.001 |
| | mmFormer [5] w/ Ours | 63.36 | 51.30 | 55.50 | 66.19 | 71.30 | 60.40 | 69.46 | 67.48 | 69.72 | 75.20 | 76.08 | 71.45 | 76.39 | 72.32 | 76.94 | 68.21 | < 0.001 |
| | M$^2$FTrans [3] | **64.96** | 53.39 | **58.16** | 64.11 | **72.58** | 62.76 | 71.78 | 68.21 | 68.78 | 74.68 | **76.48** | 72.86 | 76.21 | 73.39 | 77.20 | 69.04 | < 0.001 |
| | M$^2$FTrans [3] w/ Ours | 61.39 | 53.23 | 57.88 | 67.73 | 71.33 | 62.70 | **71.95** | **68.92** | 70.28 | 76.54 | 77.31 | **73.79** | 76.85 | **74.07** | 78.19 | 69.48 | < 0.001 |
| | IMS$^2$Trans [4] | 63.17 | 45.99 | 54.57 | 65.43 | 72.28 | 58.39 | 68.38 | 66.10 | 69.12 | 75.49 | 75.97 | 70.64 | 76.74 | 73.16 | 77.35 | 67.52 | < 0.001 |
| | IMS$^2$Trans [4] w/ 3CM | 63.14 | 44.85 | 55.09 | 67.05 | 72.01 | 58.31 | 68.82 | 65.81 | 69.32 | 75.34 | 75.73 | 70.47 | 76.51 | 72.69 | 76.73 | 67.46 | < 0.001 |
| | IMS$^2$Trans [4] w/ Ours | 63.45 | 45.07 | 53.96 | 66.91 | 71.49 | 58.11 | 69.70 | 66.44 | 70.60 | 75.39 | 76.53 | 71.67 | 76.98 | 72.85 | 77.78 | 67.80 | < 0.001 |

Table A5: **Comparison of our framework with state-of-the-art methods trained on a larger region-of-interest (ROI) size.** The average Dice Similarity Coefficient (DSC, %) and 95th percentile Hausdorff Distance (HD95, mm) were reported across five tumor subregions: whole tumor (WT), tumor core (TC), enhancing tumor (ET), necrotic and non-enhancing tumor (NCR/NET), and edema (ED). Performance was evaluated at ROI sizes of 64 and 80, with the latter being widely adopted by state-of-the-art.

| **ROI=64** | | | | | | | | | | |
|---|---|---|---|---|---|---|---|---|---|---|
| Model | Avg DSC (%) | | | | | Avg HD95 (mm) | | | | |
| | WT | TC | ET | NCR/Net | ED | WT | TC | ET | NCR/Net | ED |
| mmFormer [5] | 83.48 | 74.24 | 58.42 | 55.98 | 67.29 | 17.42 | 20.75 | 13.84 | 15.78 | 13.86 |
| mmFormer [5] w/ Ours | 84.51 | 74.46 | 59.23 | 56.46 | 68.21 | <u>10.82</u> | 14.72 | 9.42 | 11.22 | **8.89** |
| $M^2$FTrans [3] | 84.54 | 75.26 | 59.46 | 56.76 | 69.04 | 17.72 | 22.48 | 16.92 | 16.56 | 12.64 |
| $M^2$FTrans [3] w/ Ours | <u>85.53</u> | <u>76.83</u> | <u>61.54</u> | <u>57.74</u> | <u>69.48</u> | 11.87 | <u>11.33</u> | <u>6.98</u> | <u>10.07</u> | 11.01 |
| **ROI=80** | | | | | | | | | | |
| mmFormer [5] | 82.60 | 75.94 | 60.86 | 56.21 | 65.99 | 15.52 | 14.10 | 8.39 | 14.19 | 14.63 |
| $M^2$FTrans [3] | **86.64** | **78.60** | **66.74** | **58.26** | **69.75** | **8.90** | **8.36** | **4.94** | **9.50** | <u>9.05</u> |
